# Psoriasis is associated with elevated gut IL-1α and intestinal microbiome alterations: results of a cross-sectional study from Central Asia

**DOI:** 10.1101/2020.05.16.20103978

**Authors:** Sergey Yegorov, Dmitriy Babenko, Samat Kozhakhmetov, Lyudmila Akhmaltdinova, Irina Kadyrova, Ayaulym Nurgozhina, Madiyar Nurgaziyev, Sara V. Good, Gonzalo H. Hortelano, Bakytgul Yermekbayeva, Almagul Kushugulova

**Affiliations:** School of Science and Humanities, Nazarbayev University, Nur-Sultan, Kazakhstan; Faculty of Education and Humanities, Suleyman Demirel University, Almaty, Kazakhstan; Karaganda Medical University Research Centre, Karaganda, Kazakhstan; Laboratory of Human Microbiome and Longevity, National Laboratory Astana, Nazarbayev University, Nur-Sultan, Kazakhstan; Department of Biology, University of Winnipeg, Winnipeg, Canada; University Medical Center Corporate Fund, Nur-Sultan, Kazakhstan

**Keywords:** Psoriasis, gut health, cytokines, immunoglobulins, microbiome, Kazakhstan, Central Asia

## Abstract

**Objective:** Psoriasis is a chronic inflammatory condition that predominantly affects the skin and is associated with extracutaneous disorders, such as inflammatory bowel disease and arthritis. Changes in gut immunology and microbiota are important drivers of proinflammatory disorders and could play a role in the pathogenesis of psoriasis. Therefore, we explored whether psoriasis in a Central Asian cohort is associated with alterations in select immunological markers and/or microbiota of the gut.

**Setting:** We assessed correlates of psoriasis in a community from Kazakhstan.

**Participants:** Outpatients, aged 30–45 years, of a dermatology clinic presenting with plaque, guttate or palmoplantar psoriasis (n=20), and age-sex matched subjects without psoriasis (n=20).

**Design:** We undertook a cross-sectional study of stool samples. Stool supernatant was subjected to multiplex ELISA to assess the concentration of 47 cytokines and immunoglobulins and to 16S rRNA gene sequencing to characterize microbial diversity in both psoriasis+ participants and controls.

**Results:** The psoriasis+ group tended to have higher concentrations of most analytes in stool (29/47=61.7%) and gut IL-1α was significantly elevated (4.19-fold, p=0.007) compared to controls. Psoriasis was associated with alterations in gut *Firmicutes*, including elevated *Faecalibacterium* and decreased *Oscillibacter* and *Roseburia* abundance, but no association was observed between gut microbial diversity or *Firmicutes/Bacteroidetes* ratios and disease status.

**Conclusions:** Psoriasis may be associated with gut inflammation and dysbiosis. Studies are warranted to explore the use of gut microbiome-focused therapies in the management of psoriasis in this under-studied population.

## INTRODUCTION

Psoriasis is a chronic autoimmune condition that predominantly affects the skin and manifests with variable severity^1^. The clinical classification of psoriasis is based on the pattern and extent of cutaneous involvement. For example, the most common phenotype, psoriasis vulgaris, or plaque-type psoriasis, is distinguished by the presence of well-defined areas of erythematous and indurated plaques and infiltration of the epidermis and dermis by mononuclear cells^1^. Frequently, psoriasis is associated with extracutaneous manifestations, such as psoriatic arthritis (PsA) or cardiovascular disease, indicating that systemic inflammatory processes likely underlie psoriatic disease^1 2^.

The causes of psoriasis are incompletely understood; genetic predisposition plays a major role^1^, but other factors such as systemic inflammation and microbiota alterations have also been implicated in disease pathogenesis. Furthermore, the link between psoriasis and inflammatory bowel disease^3^ intriguingly points at the gut as an important contributor to psoriasis development. In support of this, recent studies have reported major alterations in both gut microbial communities^4–7^ and markers of immune response^6^ in individuals with psoriasis.

In earlier work, we characterized the gut microbiome of adults with and without metabolic syndrome from Kazakhstan, a country in Central Asia^8^. As part of our overarching objective to better understand the relationship between the mucosal and systemic correlates of chronic disease in this region^8 9^, here we expand on our earlier findings and focus on the gut microenvironment in individuals with psoriasis. We hypothesized that psoriasis in adult Central Asians is associated with gut inflammation and microbiome alterations. To test this hypothesis, we assessed levels of select cytokines and immunoglobulins and performed a metagenomic analysis of stool samples obtained from dermatology clinic outpatients in the capital city of Kazakhstan.

## METHODS

### Study setting and participant recruitment

Participants aged 30–45 years were recruited through an outpatient dermatology clinic at the Centre for Dermatology and STD prophylaxis in the capital city of Kazakhstan, Nur-Sultan (formerly Astana). The dermatological assessment of participants was done in accordance with the national clinical guidelines for psoriasis diagnosis and treatment of Kazakhstan. Study exclusion criteria were: use of antibiotics within three months prior to the study, a diagnosis of psoriatic arthritis, presence of any other chronic condition of the skin or gastrointestinal tract and presence of any severe comorbidity, or pregnancy. Healthy controls were recruited from the local communities through community-wide advertisement of the study and matched by age, sex and ethnicity. Fecal samples from all study participants were frozen within 2 hours of collection and kept frozen at –80C until analysis. This study was designed as an exploratory study to supply pilot data for future studies in the same population, therefore no formal sample size calculations were performed and the sample size was determined based on the available study budget; in total forty individuals were included in the study (n=20 with psoriasis, n=20 controls). All experimental assays were performed by research personnel blinded to the psoriasis status of participants. All research procedures were approved by the institutional review board of the UMC CF Academic Council. Written consent to participate was collected from all participants. We used the STROBE cross sectional checklist when writing our report^10^.

### Cytokine measurements

Frozen stool samples were thawed, and approximately 4 mg of each sample were dissolved in 200 μl of phosphate buffered saline. The supernatant obtained by centrifugation at 16,000 g for 15 min was then analyzed using the Milliplex Map Human Magnetic Bead Panels for cytokines and chemokines (HCYTMAG-60K-PX41) and immunoglobulins (HGAMMAG-301K-06) according to the manufacturer’s protocol on a Bio-Plex 3D instrument (Bio-Rad). The ELISA assay details are given in **Supplementary Table 1**.

### Microbiome analysis

***DNA extraction and sequencing***. A subset of 21 stool samples (14 psoriasis+ and 7 controls) was available for microbiome analysis. Samples were thawed prior to DNA extraction using QIAamp DNA Mini Kit (Qiagen). The quality of extracted DNA was assessed using Qubit dsDNA HS Assay Kit (Thermo Fisher) on a Qubit 2.0 according to the manufacturer’s manual (Invitrogen, Life Technologies). Next-generation sequencing libraries were prepared with NEXTflex 16S V1-V3 Amplicon-Seq Kit (PerkinElmer), and library quality assessed using the Qubit 2.0 system. Amplicons (96 samples per lane) were sequenced using the MiSeq platform (Illumina). ***Sequence analysis***. The QC and raw sequence pre-processing were performed using fastp (v. 0.20.0. April 2019)^11^ with the following parameters: mean quality for 4 bp window size was 20, the adapter detection enabled, reads less than 120 bp were discarded, unmerged reads were included in the final FASTQ files. Total Sum Scaling (TSS) per-sample normalization was used to remove technical bias related to different sequencing depths in among libraries and then scaled to units of reads per million per library. Taxonomic assignment was done using the naive Bayesian classifier method as implemented in the dada2 Bioconductor R package using an RDP training set (v.16).

### Statistical analysis

All statistical analyses and graphing were performed using IBM SPSS V.23 (NY, US) and GraphPad Prism V.6.0. (CA, US), unless specified otherwise. Differences in demographic characteristics between groups were assessed using Independent-Samples Mann-Whitney U and Chi-Square Tests. An ELISA analyte was considered “detectable” if the measured analyte concentration was equal to or above the assay’s lowest level of detection (LLOD), and “undetectable” if the measured concentration was below the LLOD (**Supplementary Table 1**). Analytes were grouped into i) “detectable in ≤50% of the participants” and analyzed as continuous variables using Independent-Samples Mann-Whitney U or ii) “detectable in ≤50% of the participants” and analyzed as dichotomous variables. and The α and β microbial diversities were estimated using Shannon’s diversity index and UniFrac weighted distance with Principal Coordinate Analysis (PCoA), respectively. Group comparisons for β-diversity were done via permutational multivariate ANOVA (PERMANOVA) of dissimilarity using adonis2 function from the vegan R package^12^. The linear discriminant analysis (LDA) effect size (LEfSe) algorithm was applied to identify features significantly different between the comparison groups with default settings (p≤0.05 based on Kruskal-Wallis test and LDA score≥2)^13^. Statistical analysis of microbiome data was performed using MicrobiomeAnalystR package^14^. Pearson correlation analysis between the OTU relative abundance, PASI scores and IL1α were performed on Log10-transformed data. Data with missing values were excluded from the analyses.

### Patient and Public Involvement

Patients or the public were not involved in the design, conduct, or reporting of this research.

## RESULTS

### Participant demographics and clinical characteristics

A summary of the socio-demographic characteristics and psoriasis diagnostic data are given in **Table 1**. Age, sex or BMI did not significantly differ between groups. The median participant age was 33 years and significantly more individuals were not married in the psoriasis group (7/20) compared to the controls (1/20, p=0.018). In the majority (85%) of the participants, one or both parents had psoriasis. Psoriasis vulgaris was most prevalent (75%) while 25% of the participants had guttate and palmoplantar psoriasis. Most participants (85%, 17/20) reported having had the condition for > 5 years, indicating seasonality in symptom presentation. In most participants psoriasis was associated with hair, but not nail, damage. The median psoriasis area and severity index (PASI) score was 11.4; mild, moderate and severe forms^15^ were seen in 25, 35 and 40% of the participants, respectively (**Table 1**).

**Table 1.** Socio-demographic and psoriasis-specific characteristics of participants.

| Participant characteristic | Psoriasis group (N=20) | Controls (N=20) | P value |
| --- | --- | --- | --- |
| **Median age (IQR)** | 34.5 (31.0-37.8) | 33.0 (31.3-34.0) | 0.205 |
| **Men, n (%)** | 10 (50.0) | 11 (55.0) | **0.752** |
| **Mean body mass index (range)^** | 24.8 (21.4-28.7) | 23.9 (18.6-32.7) | 0.475 |
| **Married, n (%)** | 13 (65.0) | 19 (95.0) | **0.018** |
| **Psoriasis type** |  |  |  |
| Vulgaris | 15 (75.0) | - | - |
| Guttate | 3 (15.0) | - | - |
| Palmoplantar | 2 (10.0) | - | - |
| **Psoriasis present in a parent** | 17 (85.0) | - | - |
| **Time since psoriasis first noted** |  |  |  |
| <5 years | 3 (15.0) | - | - |
| 5-10 years | 9 (45.0) | - | - |
| >10 years | 8 (40.0) | - | - |
| **Head hair damage present** | 17 (85.0) | - | - |
| **Nail damage present** | 1 (5.0) | - | - |
| **Median PASI (IQR)** | 11.4 (6.7-16.4) | - | - |
| **Psoriasis severity based on PASI** |  |  |  |
| Mild [PASI<7], n (%) | 5 (25.0) | - | - |
| Moderate [7≤PASI≤12], n (%) | 7 (35.0) | - | - |
| Severe [PASI>12], n (%) | 8 (40.0) | - | - |

BMI, body mass index; IQR, interquartile range; PASI, Psoriasis Area and Severity Index

### Comparison of stool-derived cytokine profiles

Samples from a total of 40 participants were analyzed. Cytokine and immunoglobulin data were obtained for 22 and 29 (out of a total of 40) participants, respectively. Repeated efforts to obtain ELISA data for outstanding measurements were unsuccessful, likely due to sample characteristics that were psoriasis-independent. Overall, 26/41 cytokines and chemokines and 3/6 Igs were detectable (i.e. >0 pg/ml in >50% of the participants); the psoriasis+ group tended to have higher median concentrations for most analytes (29/47=61.7%) compared to the control group (**Supplementary Table 2**); no correlation was observed between the stool analytes and the PASI scores. IL-1α was the only cytokine significantly elevated in the psoriasis+ group (4.19-fold, p=0.007, **Supplementary Table 2** and **Figure 1a**); this difference was not associated with differences in socio-demographic or psoriasis-specific characteristics in this subset of participants (N=19, **Supplementary Table 3**).

**Figure 1.**
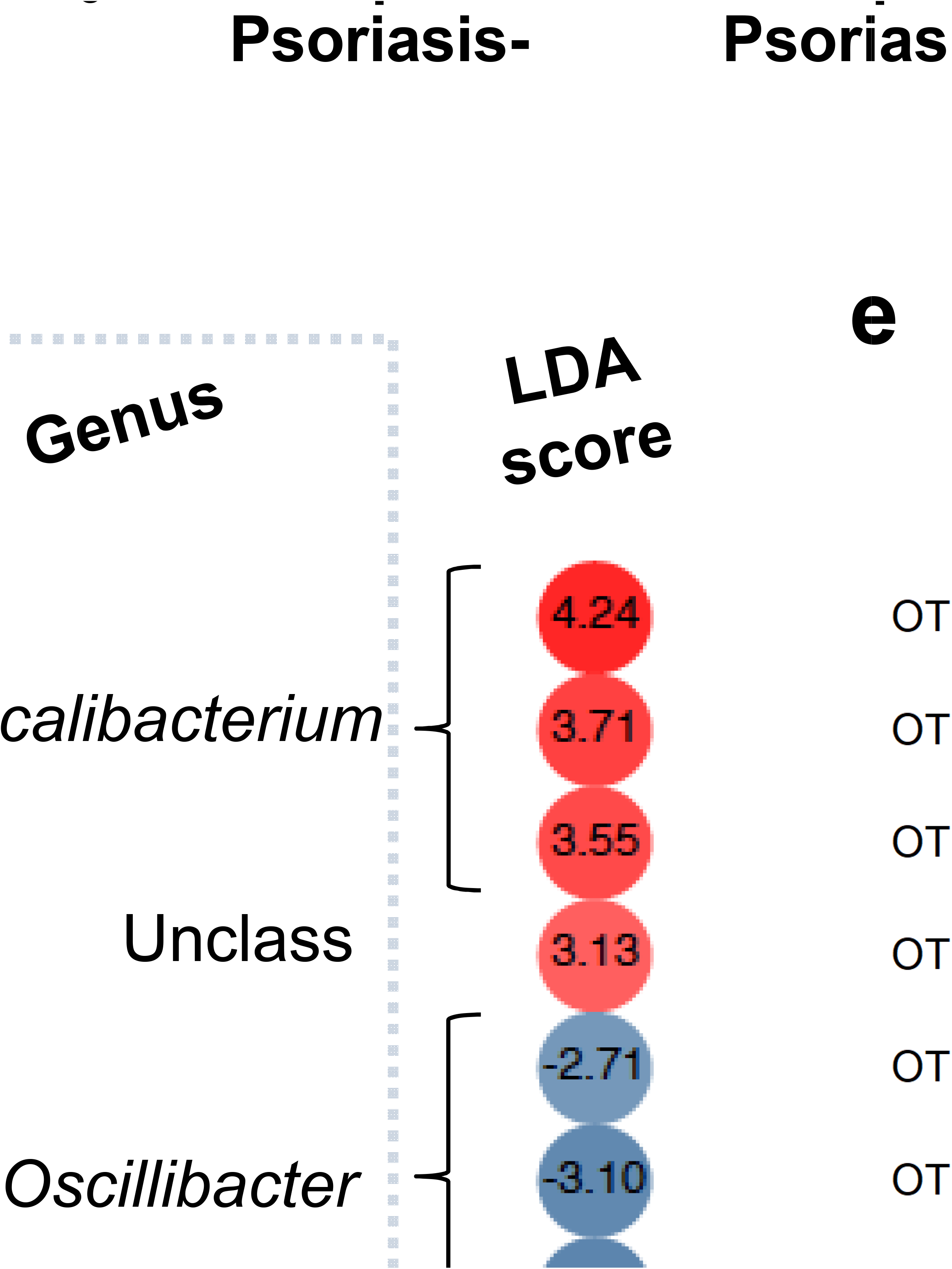
Gut IL-1α and microbiata differences between psoriasis+ and control participants. **a**. Gut IL-11α concentration of psoriasis+ individuals and controls. The IL-1 data were available for 23/40 (psoriasis+, N=13, and psoriasis-, N=10) participants. **b**. Microbial species diversity measured by Shannon index in the gut of psoriasis+ individuals and controls. **c**. Gut *Firmicutes/Bacteroides* ratio of psoriasis+ individuals and controls. **d**. Heatmap showing the LefSe-derived LDA scores for each of the OTU (n=18) differentially abundant in the gut of psoriasis+ participants. Data based on N=21 samples. **e**. Heatmap based on the Pearson coefficients of correlation between the psoriasis-associated microbial taxa, IL-1α and PASI scores. Analysis based on N=14 samples. Significant correlations marked by asterisks (*).

### Comparison of gut microbiome profiles

#### Gut microbiota diversity and Firmicutes/Bacteroides (F/B) ratio

A total of 21 fecal samples, 14 psoriasis+ and 7 controls, were available for the microbiome analysis. When compared to controls, neither the microbial diversity indices nor the *F/B* ratios of the psoriasis patients were significantly different (**Figure 1b** and **Figure 1c**, and **Supplementary Figure 1**), although the median *F/B* ratio in the psoriasis group was double of that observed in the controls (2.5 vs 1.3), largely driven by an elevated F/B ratio in three psoriasis+ participants (**Figure 1c**).

#### Bacterial taxa associated with psoriasis

The LefSe analysis (12) identified 18 operational taxonomic units (OTUs) that could significantly discriminate among groups at p=0.05 (**Figure 1d**). After correcting for multiple hypothesis testing, no OTUs remained significantly different between groups (FDR=0.05); notwithstanding the overall differences in bacterial composition observed here are in agreement with those observed in other studies^5 6^. Specifically, all 18 OTUs that were found to be differentially abundant were from the phylum *Firmicutes* and, except one unclassified OTU, were classified as *Clostridia*, Order *Clostridiales*. The majority of these OTUs (14/18, 77.8%) were less abundant in psoriasis+ participants compared to controls. At the family level, all 8 OTUs from the Lachnospiraceae were found to be less abundant in psoriasis+ patients, while 4 of the 7 OTUs from the family *Ruminococcaceae* were more abundant in psoriasis+ patients and 3 were less abundant (Figure 1d). Examining this at the level of genus indicates that all OTUs from *Faecalibacterium* (n=3 OTU) exhibited increased abundance, while those from the *Oscillibacter* (n=3 OTU) and *Roseburia* (n=1 OTU) exhibited reduced abundance in the psoriasis group (**Figure 1d**). Lastly, the correlation analyses confirmed that there was a negative correlation between OTU abundance and both disease severity (PASI) and IL-1α levels for most OTUs; only two of these correlations reached statistical significance (Figure 1e). Three taxa, OTU_0131 (from the genus *Faecalibacterium*), OTU_1333 (unclassified) and OUT_1481 (unclassified) exhibited positive correlations between OTU abundance and PASI and IL-1α. (**Figure 1e**).

## DISCUSSION

Here, we examined associations between psoriasis, and gut immunology and microbial parameters in an adult cohort based in Kazakhstan. We found that psoriasis was associated with elevated gut IL-1α and altered abundance of *Firmicutes*. Overall, these findings are consistent with the notion that psoriasis is linked to gut dysbiosis and inflammation^5 6 16 17^.

Few studies to date have directly assessed the immunological changes occurring in the intestine of psoriasis patients. Scher and co-authors reported elevation of soluble IgA and reduction of receptor activator of nuclear factor kappa-B ligand (RANKL), a critical factor controlling differentiation of intestinal lamina propria cells, in PsA^6^. In the current study, we saw trends toward elevation for >60% of the analytes from the stools of psoriatic individuals, although only IL-1α, a cytokine central to the regulation of inflammation^18^, was significantly elevated compared to the controls. Elevated cytokines in stool were previously detected in the context of gut inflammation^19 20^, while in a murine model of colitis IL-1α secreted by intestinal epithelium was the main driver of immune activation^21^. In psoriasis, IL-1α drives the formation of dermal clusters of T cells and antigen presenting cells and is involved in the development of dermal Th17 responses^22–24^. Therefore, it is possible that increased levels of gut IL-1α in individuals with psoriasis may contribute to increased inflammation via the gut-skin axis^25^ and may help explain the well-documented epidemiological link between psoriasis and inflammatory bowel diseases^3^.

To the best of our knowledge this study is the first to characterize psoriasis in a population from Kazakhstan and to assess its associations with gut immunology and microbiome. Earlier, we performed a large-scale metagenomic analysis and compared the Kazakh gut metagenome to its counterparts from other regions of the world^8^. We found significant differences between the microbiomes of Kazakhs and both Europeans and East Asians. One distinguishing feature of the Kazakh gut metagenome is a remarkably dominant *Prevotella*-rich enterotype, which is typically regarded as proinflammatory^26^, and was associated with the metabolic syndrome in our cohort^8^. In our current analysis, psoriasis was not associated with changes in gut *Prevotella* abundance, but, consistent with studies from other cohorts, was associated with alterations in *Firmicutes*^5 6^. Although the direction of *Firmicute* change differs among studies, this suggests that psoriasis-associated gut microbiome signatures are generalizable to human populations residing in different parts of the world, but may are modified by differences in environmental factors and diet.

We found that psoriasis was associated with an elevated abundance of members of the genus *Faecalibacterium*, which is consistent with a metagenomic analysis from Spain^5^. On the other hand, a study based on a PCR-aided identification of specific bacterial species in the Netherlands found that stool samples from psoriasis patients were depleted for *Faecalibacterium prausnitzii*^4^. Notably, gut *Faecalibacterium* alterations have been associated with eczema and IBD^27 28^ and, taken together, these data highlight the dynamic role gut *Faecalibacterium* spp. are potentially playing in the pathogenesis of both skin- and gut diseases.

The causality of the relationship between psoriasis and gut immunology and microbiota is unclear. Psoriasis is associated with systemic inflammation^1^, which could drive changes in the gut mucosa and thereby affect the gut microbiota. It has also been proposed that the proinflammatory environment of the gut, precipitated by dietary insults and genetic predisposition, could induce systemic inflammation and thus cause skin-directed inflammation^29^. Future mechanistic studies may clarify the exact mechanisms involved, meanwhile our finding of enhanced IL-1α, a key regulator of inflammatory processes, suggests that the gut may play a central role in the induction of inflammatory responses in psoriasis^18^. While current therapies for mild psoriasis target cutaneous manifestations and exhibit relatively low efficacies, the more efficacious therapeutic modes for severe psoriasis based on the use of antibodies are expensive and can have severe side effects^1^. If the psoriatic disease is mediated by the gut immune milieu and microbiota, psoriasis treatment could benefit from modulating the gut microenvironment using microecologic agents, such as probiotics, and fecal microbiota transplantation.

Our findings should be interpreted in the light of several limitations. First, due to technical and logistic limitations our study had a small sample size and both the ELISA and metagenomic analysis results were only available for subsets of the original cohort. Second, soluble analytes were measured in stool supernatant, a highly heterogeneous sample, and further improvements of the protocol may improve cytokine yields; work is underway in our laboratory in this direction. Lastly, this study used an exploratory approach and therefore follow-up studies will be required to confirm these findings. It is noteworthy that despite age and sex-matching, there were significant differences in the marital status of psoriasis+ participants and controls. Psoriasis is a socially stigmatizing condition that affects the individual’s ability to build relationships^1^, underscoring the need for better therapeutic and psychological instruments to help individuals and their families affected by psoriasis.

In summary, this study for the first time assessed psoriasis associations with gut immunology and microbiome in an under-studied population from Central Asia. Although limited by a small sample size, the data presented here provide more evidence in support of the notion that gut immunology and microbiota may play critical roles in the pathogenesis of psoriasis, an autoimmune condition that is commonly thought to primarily affect the skin. Future studies are warranted to explore the inclusion of gut microenvironment modulating agents in the treatment and prophylaxis of psoriasis in this population.

## Data Availability

All processed ELISA and microbiome data generated and analyzed during this study are included in this published article as supplementary data. The raw microbiome sequence data will be uploaded to a public database upon article publication.

## Strengths and limitations of this study

- This is the first study to characterize psoriasis in a population from Kazakhstan and to assess its associations with gut immunology and microbiome.
- Our cross-sectional analysis of soluble immune mediators and microbiota provides insights into associations of psoriasis with gut inflammation and dysbiosis, particularly highlighting the role of interleukin-1 and *Firmicutes* in these processes.
- Our study had a small sample size and some results were only available for subsets of the original cohort.
- Soluble immune function mediators were measured in stool supernatant, known to be highly heterogeneous, and further improvements of the protocol may improve analyte yields; work is underway in our laboratory in this direction.

## DECLARATIONS

### Acknowledgements

We thank all the participants and research teams involved in the study.

### Author contributions

Conceptualization, SY, DB, SVG, BY, AK; Data curation, SY, DB, SVG, BY, AK; Formal analysis, SY, DB, SVG; Funding acquisition, BY; Methodology, SK, LA, IK, AN, BY, AK; Project administration, BY, AK, GH; Resources, BY, AK, GH; Supervision, BY, AK, GH; Visualization, SY, DB, SVG; Writing – original draft, SY, DB; Writing – review & editing, all authors.

### Ethics approval and consent to participate

All study procedures were approved by the UMC ethics committee. Written informed consent was obtained from all participants.

### Supplementary information

Supplementary information for this article includes Supplementary Tables 1–3, Supplementary Figure 1 and Supplementary Dataset 1 (raw dataset).

